# Dermal microdialysis: a method to determine drug levels in the skin of patients with Post Kala-azar Dermal Leishmaniasis (PKDL)

**DOI:** 10.1101/2023.01.09.23284369

**Authors:** Gert-Jan Wijnant, Srija Moulik, Kingshuk Chatterjee, Nilay K Das, Raúl de la Flor, Katrien Van Bocxlaer, Simon L. Croft, Mitali Chatterjee

## Abstract

**Objectives:** Post-kala-azar-dermal leishmaniasis (PKDL) is an infectious skin disease that occurs after apparent cure from visceral leishmaniasis (VL) and causes stigmatizing lesions on the face and other exposed body parts. While the first-line drug miltefosine is typically used for 28 days to treat VL, 12 weeks of therapy is required for PKDL, highlighting the need to evaluate the extent of drug penetration at the dermal site of infection. In this proof-of-concept study, we demonstrate the use of a minimally invasive sampling technique called microdialysis to measure dermal drug exposure in a PKDL patient, providing a tool for the optimisation of treatment regimens.

**Methods and Materials:** Three PKDL patients receiving miltefosine (50 mg twice daily for 12 weeks) were recruited to the study. Conventional blood samples and skin biopsies were collected to determine drug content. One patient consented to undergo dermal microdialysis. Briefly, a µDialysis Linear Catheter 66 for skin and muscle, a probe with a semi-permeable membrane, was inserted in the dermis. A perfusate (a drug-free physiological solution) was pumped through the probe at a low flow rate, allowing miltefosine present in the dermis to cross the membrane and be collected in the dialysates over time. Protein-free (dialysates) and total (blood and skin biopsies) drug concentrations were analysed using LC-MS/MS.

**Results and conclusions:** After confirming the presence of miltefosine in skin lesions of PKDL patients via traditional biopsy sampling, protein-free drug concentrations over time (C_max_ ≈ 1 µg/ml) could be detected in the infected dermis via microdialysis. This clinical proof-of-concept study thus illustrates the potential of dermal microdialysis as a minimally invasive alternative to invasive skin biopsies to quantify drug concentrations directly at the pharmacological site of action in PKDL.

## Introduction

Post-kala azar dermal leishmaniasis (PKDL) is a dermal complication that follows apparent cure from visceral leishmaniasis (VL), a fatal disease caused by *Leishmania donovani* parasites. In East Africa, PKDL occurs in up to 50% of patients, causing mostly papular lesions that tend to resolve spontaneously. In South-East Asia, the disease affects 5-10% of apparently cured VL patients and results in widespread macular or polymorphic lesions that are challenging to treat. ^[1]^ The presence of parasites in the skin form an important reservoir for the continued transmission of VL via sand fly bites, making early diagnosis and therapy critical to the success of the ongoing leishmaniasis elimination programme. ^[2,3]^ However, the existing chemotherapeutic options show variable cure rates, require hospital administration (intravenous infusion of liposomal amphotericin B), or suffer from poor compliance due to long treatment durations and frequent side effects (oral miltefosine, MF). ^[4]^

To date, pharmacokinetics (PK) have not been used to support the optimal design of drug dose regimens to treat PKDL, which remain empirical. ^[5]^ Measuring drug concentrations in skin tissue, the PKDL target site, is challenging due to the invasive nature of traditional sampling methods (skin biopsies) and the complexity of the associated sample preparation. ^[6]^ An attractive alternative to studying skin PK is dermal microdialysis (DMD), a minimally invasive sampling method that relies on a small probe, a semi-permeable membrane, that is temporarily implanted in the dermis. Upon slow perfusion of the probe with a physiological buffer, the protein-free drug concentrations can be recovered from the dermal extracellular fluid and measured in the dialysates. ^[7,8]^ The technique has already been successfully applied for clinical PK studies in various dermatoses, including psoriasis and atopic dermatitis, but not in PKDL. ^[9]^ In this article, we report a proof-of-concept study investigating DMD to measure MF exposure in the skin of a PKDL patient, describing the methodology and outcome of this approach as a basis for future dermato-pharmacokinetic investigations to facilitate the design of better treatment regimens and combinations.

## Material and Methods

Three patients were recruited with a confirmed diagnosis of PKDL (qPCR positive) ^[10]^ and received oral MF therapy (Impavido® 50 mg capsules twice daily for 12 weeks, Table 1). Before performing the microdialysis procedure, conventional PK sampling methods were applied to confirm the presence of MF in systemic circulation (heparinised blood) and lesional skin (4 mm biopsy, 5-20 mg weight). MF was extracted from plasma and skin homogenates using acetonitrile (1:5 ratio) and drug levels determined using a validated LC-MS/MS method with a lower limit of detection of 2 ng/ml (Supplementary Material 1).

**Table 1:** Study population details

|  | **Patient 1** | **Patient 2** | **Patient 3** |
| --- | --- | --- | --- |
| **Gender** | Male | Male | Female |
| **Age (in years)** | 45-50 | 35-40 | 15-20 |
| **History of VL** | No | Yes | Yes |
| **PKDL lesion characteristics** | Macular (back, shoulders) | Macular (back, shoulders, arms) | Macular lesions (back, face) |
| **Lesion sampling site** | Lower back | Lower right arm | Lower back |
| **Days on MF treatment** | 43/84 | 72/84 | 54/84 |
| **Parasite load at start of treatment (number of parasites per $\mu\text{g}$/genomic DNA)** | 12,582 | 855,452 | Not available |
| **Parasite load during MF treatment (number of parasites per $\mu\text{g}$/genomic DNA)** | 8,568 (- 31.9 %) | 6,654 (- 99.2 %) | 512 |
| **Total drug in plasma ($\mu\text{g}/\text{ml}$)** | 3.072 | 5.778 | 0.124 |
| **Total drug in lesion ($\mu\text{g}/\text{g}$)** | 3.389 | 3.298 | 0.131 |

The optimal experimental conditions to sample and recover MF were determined *in vitro*. Briefly, the probe of choice, a µDialysis Linear Catheter 66 for skin and muscle (20 kDa pore cut-off size, 10 mm length, mDialysis, Sweden), was flushed with different perfusate compositions at various flow rates and immersed in a reference solution containing 100 ng/ml MF in Ringer’s solution (at 34°C and under slow, continuous stirring to mimic the dermal micro-environment). Dialysates containing recovered MF were collected every 30 minutes in glass vials containing acetonitrile (1 volume solvent per 3 volumes sample) and the drug content was analysed via LC-MS/MS (supplement 1). The *in vitro* relative recovery value (RR) for MF was calculated as the ratio of the average recovered drug concentration in the dialysates over the drug concentration in the reference solution containing the probe (supplement 2). The DMD conditions that resulted in the highest *in vitro* RR value (35.5%) consisted of a flow rate of 1 µl/min, and Ringers physiological buffer with 5% β-cyclodextrin (w/v) as perfusate composition. β-cyclodextrin was added to the perfusate to enhance the aqueous solubility of MF and reduce aspecific binding of the lipophilic compound to the polymeric microdialysis tubing. This perfusate was prepared under sterile conditions and did not cause any adverse effects upon prior administration in healthy volunteers via microdialysis (data not shown).

One patient (patient 1, Table 1) consented to undergo the DMD procedure (Figure 1b) on day 43 of MF treatment (Figures 1a and 1b). Under local anaesthesia (0.1 ml lidocaine hydrochloride 2%, ID), microdialysis probes were intradermally inserted into a macular PKDL lesion and a nearby unaffected skin site on the back of the patient. Probes were perfused as described above. After allowing the skin to recover from the insertion trauma for 1 hour, a tablet containing 50 mg oral MF was administered and dialysates were collected every 30 minutes for LC/LC-MS analysis. The detected miltefosine concentrations were corrected using the *in vitro* RR value. No adverse effects were observed during DMD or the weeks following the procedure.

**Figure 1.**
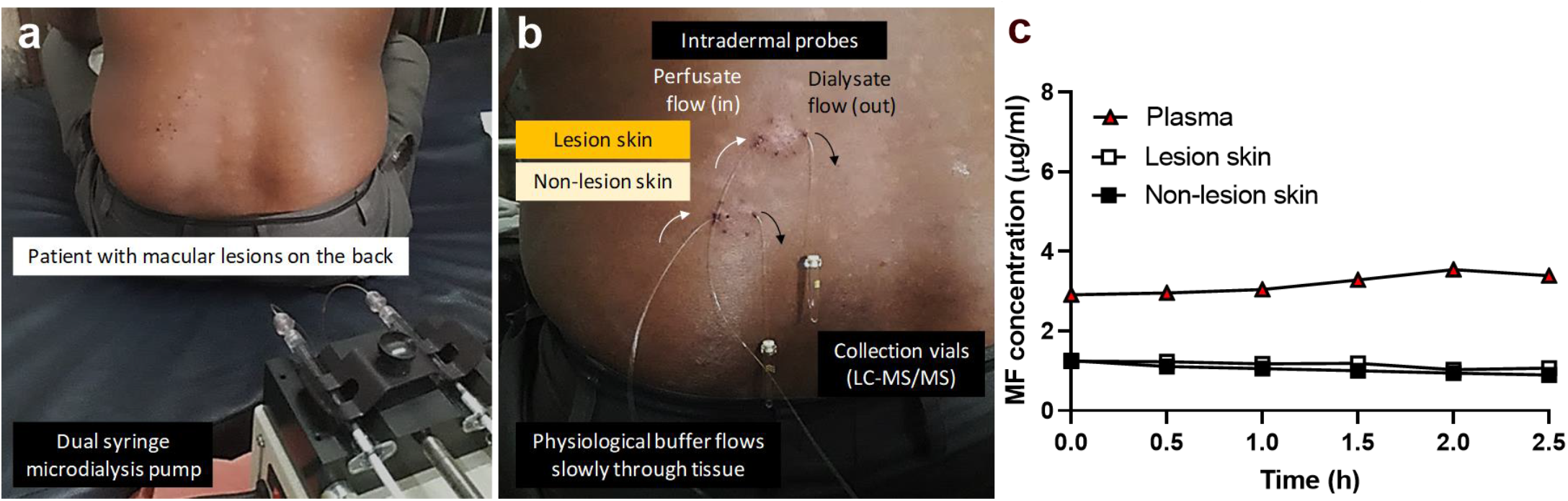
Validation of dermal microdialysis as a method to measure drug exposure in the skin of a PKDL patient receiving miltefosine (MF). (a) Patient of PKDL with macular lesions on the back at the start of the procedure. (b) Experimental set-up of the microdialysis technique, with linear catheters (20 kDa pore cut-off) inserted intradermally in a macular skin lesion and a non-lesional skin site. Using a dual syringe microdialysis pump, the microdialysis catheters were perfused at a flow rate of 1 µl/min with Ringer’s physiological solution containing 5% β-cyclodextrin, allowing recovery of MF from the dermis and LC-MS/MS quantification in the dialysates. (c) Pharmacokinetics (PK) of MF after oral administration of 1 × 50 mg miltefosine (Impavido®) capsule to a PKDL patient on day 43/84 of treatment. Plasma levels (▲) represent total (protein-free + protein-bound) drug concentrations derived from whole blood samples. Lesional (□) and non-lesional (■) skin levels represent protein-free drug concentrations sampled by dermal microdialysis, corrected for the in vitro relative recovery value (35.7%).

## Results

In all patients, MF was detected in plasma and skin (Table 1), concordant with published data showing extensive plasma and tissue accumulation upon repeated oral dosing.^[11]^ Compared to patients 1 and 2, patient 3 showed a > 20-fold reduction in systemic and skin concentrations despite similar treatment duration, potentially attributable to variations in PK depending on the type of lesion, ^[12]^ compliance, sampling and/or processing. As 43/84 days of treatment were completed, the dermal parasite load was decreased as compared to levels at presentation (Table 1).

The analysis of the microdialysis dialysates demonstrated a stable concentration of protein-free MF (Figure 1c) around 1 µg/ml for both lesional and non-lesional skin in the 2.5 hours after dosing. These skin PK results for MF appear physiologically feasible considering the drugs (i) long gastro-intestinal absorption time (up to 8 hours) (ii) extensive tissue accumulation over time and (iii) high plasma protein binding (up to 98%).^[13]^ Interestingly, while our results indicate no difference in drug penetration in macular PKDL lesions and non-lesional skin, earlier DMD studies revealed higher methotrexate bioavailability in psoriasis plaques than in non-lesional skin.^[14]^

In conclusion, this clinical proof-of-concept study has confirmed DMD, a minimally invasive sampling technique, as an attractive alternative to invasive skin biopsies to measure pharmacologically active drug concentrations in skin lesions of PKDL patients. Despite the limitations of this study, including: (i) a lack of comparative protein-free drug concentration data from skin biopsy and plasma samples, (ii) the unknown matrix effect of dermal fluid on LC-MS readouts due to *in vitro* probe calibration, (iii) the absence of an internal standard for LC-MS and (iv) the low number of study subjects (n=1) and clinical presentations (macular PKDL only), the work provides a set of methodologies that paves the way for a full investigation of the dermal PK of MF and other anti-leishmanial drugs to facilitate optimization of treatment regimens for PKDL and other forms of tegumentary leishmaniasis, especially aimed at reducing the unacceptably long duration of treatment..

## Data Availability

All data produced in the present study are available upon reasonable request to the authors

## Ethics statement

This clinical proof-of-concept study received approval from the ethics committee of the London School of Hygiene & Tropical Medicine (LSHTM), UK and Calcutta School of Tropical Medicine (CSTM), Kolkata, India under references LSHTM-15974 and CREC-STEM/2018-AS20 respectively. Patients provided written consent for participation in the study.

## Acknowledgements

GJW was supported by a Royal Society of Tropical Medicine and Hygiene (RSTMH) Small Grant Programme and the London School of Hygiene and Tropical Medicine (LSHTM) Doctoral Project Travelling Scholarship. KVB was supported by a fellowship awarded from the Research Council United Kingdom Grand Challenges Research Funder under grant agreement ‘A Global Network for Neglected Tropical Diseases’ number MR/P027989/1. MC received financial assistance from an Indian Council for Medical Research, Govt. of India, Grant number: 6/9-7[151]2017-ECD II.

