## Supplementary information for "Dermal microdialysis: a method to determine drug levels in the skin of patients with Post Kala-azar Dermal Leishmaniasis (PKDL)"

---

### Supplementary material

**Supplementary material 1**: Details of the analytical method for the quantification of miltefosine in plasma, skin homogenates and dermal microdialysis samples.

<table>
<tr><td>MS conditions</td><td>Agilent 1290 UHPLC/Agilent 6550 QToF<br>ESI: Positive ion electrospray<br>MS Scanning: Full scan m/z 100 – 1000, 3 spectra/s, centroid<br>miltefosine: [M+H]</td></tr>
<tr><td>UPLC conditions</td><td>Mobile Phase:<br>Mobile phase A: Water, 0.1% formic acid<br>Mobile phase B: Acetonitrile, 0.1% formic acid<br><br><table><tr><th>Time (min)</th><th>Mobile Phase B (%)</th></tr><tr><td>0</td><td>2</td></tr><tr><td>0.3</td><td>2</td></tr><tr><td>1.1</td><td>95</td></tr><tr><td>1.75</td><td>95</td></tr><tr><td>1.8</td><td>2</td></tr><tr><td>2</td><td>2</td></tr></table><br>Column: ACQUITY UPLC BEH C18 (2.1×50 mm, 1.7 µm)<br>Column temperature:  50°C<br>Flow rate: 0.4 mL/min<br>Retention time miltefosine: 1.80 min<br>Injection volume: 2ul (2ul loop fitted, overfilled)<br>Divert to waste: 0-0.6 min</td></tr>
</table>

Gert-Jan Wijnant[1], Srija Moulik[2], Kingshuk Chatterjee[3], Nilay K Das[4], Raúl de la Flor[5], Katrien Van Bocxlaer[1,6], Simon L. Croft[1*], Mitali Chatterjee[2]*

**Supplementary material 2**:

Method development and microdialysis probe calibration. To mimic the *in vivo* environment of the dermis, the microdialysis probe (µDialysis Linear Catheter 66 for skin and muscle, 20 kDa pore cut-off size, 10 mm length; mDialysis, Sweden) was immersed in a physiological buffer (full strength Ringer solution) at 34 °C under continuous, slow magnetic stirring. The catheter was connected to a microdialysis pump (Harvard apparatus 22, dual syringe channel pump) and perfused at a constant flow rate. Samples were collected at 30-minute intervals in glass vials containing one volume of acetonitrile per three volumes of dialysate (1:3) and analysed via LC-MS/MS (lower limit of quantification = 2 ng/ml). After one hour, 10 µl of a miltefosine stock was spiked into the probe-containing reservoir to achieve at a final drug concentration of 100 ng/ml ($C_{ref}$) and dialysates were collected ($C_{dialysate}$). *In vitro* relative recovery (RR, %) was defined as the ratio of the average $C_{dialysate}$ over $C_{ref}$. This value is typically lower than 100 % due to (i) incomplete equilibration between the drug concentrations in the environment surrounding the probe and those in the dialysate and (ii) the tendency of lipophilic compounds to stick to the polystyrene material of the microdialysis tubing. To improve the RR of miltefosine, which was extremely low under standard conditions (< 0.1%), the perfusate was supplemented with β-cyclodextrin to enhance the aqueous solubility of the compound. The experimental conditions that resulted in the highest recovery of miltefosine (5% β-CD, 1 µl/min flow rate) were applied during the *in vivo* microdialysis procedure and the corresponding *in vitro* RR value of 35.7% was used to correct the *in vivo* results.

| **Perfusate conditions** | | $C_{dialysate}$ **(ng/ml)** | | | $C_{ref}$ **(100 ng/ml)** | **Relative recovery (%)** |
|---|---|---|---|---|---|---|
| **β-cyclodextrin (% w/v)** | Flow rate (µl/min) | 30 min | 60 min | 90 min | | |
| 0 | 0.5 | < 2 | < 2 | < 2 | 130.4 | < 0.1 |
| 5 | 0.5 | 30.8 | 38.4 | 40.2 | 104.3 | 34.9 |
| 5 | 1 | 35.3 | 43.9 | 32.8 | 104.3 | 35.7 |
| 10 | 1 | 27.1 | 33.6 | 34.8 | 104.3 | 30.5 |
